# Prediction of Unplanned 30- day Readmission for ICU Patients with Heart Failure

**DOI:** 10.1101/2021.10.06.21264643

**Authors:** M Pishgar, J Theis, M Del Rios, A Ardati, H Anahideh, H Darabi

## Abstract

**Background:** Intensive Care Unit (ICU) readmissions in patients with Heart Failure (HF) result in a significant risk of death and financial burden for patients and healthcare systems. Prediction of at-risk patients for readmission allows for targeted interventions that reduce morbidity and mortality.

**Methods and Results:** We presented a process mining approach for the prediction of unplanned 30-day readmission of ICU patients with HF. A patient’s health records can be understood as a sequence of observations called event logs; used to discover a process model. Time information was extracted using the DREAM (Decay Replay Mining) algorithm. Demographic information and severity scores upon admission were then combined with the time information and fed to a Neural Network (NN) model to further enhance the prediction efficiency.

**Results:** By using the Medical Information Mart for Intensive Care III (MIMIC-III) dataset of 3411 ICU patients with HF, our proposed model yielded an Area Under the Receiver Operating Characteristics (AUROC) of 0.920.

**Conclusions:** The proposed approach was capable of modeling the time-related variables and incorporating the medical history of patients from prior hospital visits for prediction. Thus, our approach significantly improved the outcome prediction compared to that of other ML-based models and health calculators.

## BACKGROUND

The prevalence of Heart Failure (HF) rises over time. Approximately, 6 million American adults (age > 20) had HF between 2015 to 2018 [1]. Despite the progress made in HF therapeutics, readmission rates remain high at nearly 20% [2,3]. Excessive unplanned readmissions and subsequent waste of medical resources have had financial implications that directly affect the overall performance of the hospitals. The Hospital Readmissions Reduction Program was established by the Affordable Care Act (ACA) in 2010 to encourage hospitals to avoid readmissions by penalizing the hospitals that exceed the expected thresholds [4]. Since 2012, hospitals have been penalized over $2.5 billion by the Centers for Medicare & Medicaid Services (CMS) for exceeding the unplanned 30-day readmission rates [5]. Unplanned ICU readmission may impose a severe financial burden on both hospitals and patients [6]. Readmissions were found to be associated with increased morbidity and mortality. The mortality rate of unplanned ICU readmission ranged between 26% to 58% [7]. The ICU readmission rate had increased over time rising from 4.6% in 1989 to 6.4% in 2003 [1]. Approximately 16% of unplanned ICU readmissions occurred within 30-days of initial hospital discharge [8,9].

The Electronic Health Record (EHR) has been revolutionizing the health care decision-making processes through collecting and preserving medical data in a digital format. The use of the EHR has been allowing hospital systems to make intelligent data-informed decisions to address a wide range of problems from learning personalized prescriptions to maximizing the performance of hospitals [10,11].

Several ML and artificial intelligence techniques have been proposed to predict unplanned 30-day readmission of ICU patients with HF [11,12]. However, the results of the developed models were not quite reliable. Process mining analyzes and optimizes the sequence of events occurring during running processes, known as the event logs. The process mining approach has been used to enhance the healthcare processes [13]. However, the medical history of the patients from past hospital visits has not yet been used to predict unplanned readmission [14,15,16].

The present study aimed to introduce a novel process mining approach that incorporated the past medical history of the patients from prior hospital visits and the time information related to the variables (Time State Samples (TSS)) to predict unplanned 30-day readmissions of the ICU HF patients.

## METHODS

### Data Source and Inclusion Criteria

We used the Medical Information Mart for Intensive Care III (MIMIC-III) public database, which contained deidentified clinical data of the patients who were admitted to the Beth Israel Deaconess Medical Center in Boston, Massachusetts [17]. MIMIC-III contained 38,597 adult patients and 49,785 hospital admissions from 2001 to 2012. This database consisted of various tables such as admission information, demographics, caregiver information, lab values, charted observations, discharge summary notes, and diagnosis codes.

In order to identify HF patients, specific ICD-9 codes related to the HF diagnosis including 398.91, 402.01, 402.11, 402.91, 404.01, 404.03, 404.11, 404.13, 404.91, 404.93, 428. XX were used in this study. Patients were included if any of these ICD9 codes appeared in the most recent admission following the standard approach in the existing literature [18]. We also included HF patients who had visited any hospital at least once before the current ICU hospital visit. Patients who died in the same hospital ICU or got discharged from a hospital ICU and died later in another hospital or other parts of the same hospital were excluded.

### Variable Selection

Several variables were considered as the inputs to the model which are as follows: the admission type, the associated time of the admission, types of insurance, the discharge time, several lab measurements, various performed services, procedures, and diagnoses on the patients, and demographic information. The admission types were categorized either as planned or unplanned admission. The insurance group types were defined as Medicare, Medicaid, Private, Government, and Self-pay. Lab values typically obtained to predict HF were extracted for each patient including Blood Urea Nitrogen (BUN), Serum Creatinine, sodium ion, and pro-brain natriuretic peptide (NT-proBNP). The various types of performed services, procedures, and diagnoses were considered in the form of CPT and ICD-9-CM codes. The demographic information including age, gender, and ethnicity were used as additional variables in the model.

### Conversion of EHR into Event Logs

The proper format of the input data for the process mining is event logs. Event logs contain the sequences of events as well as the associated time at which specific events occurred, which are referred to as timestamps.

The transformation of the EHR of the patients into the event logs was done based on the method reported by Theis et al. [16].

Thirteen different event types were defined in this section. Table 1 shows the mapping of each of the considered event types with the MIMIC-III tables.

**Table 1:**
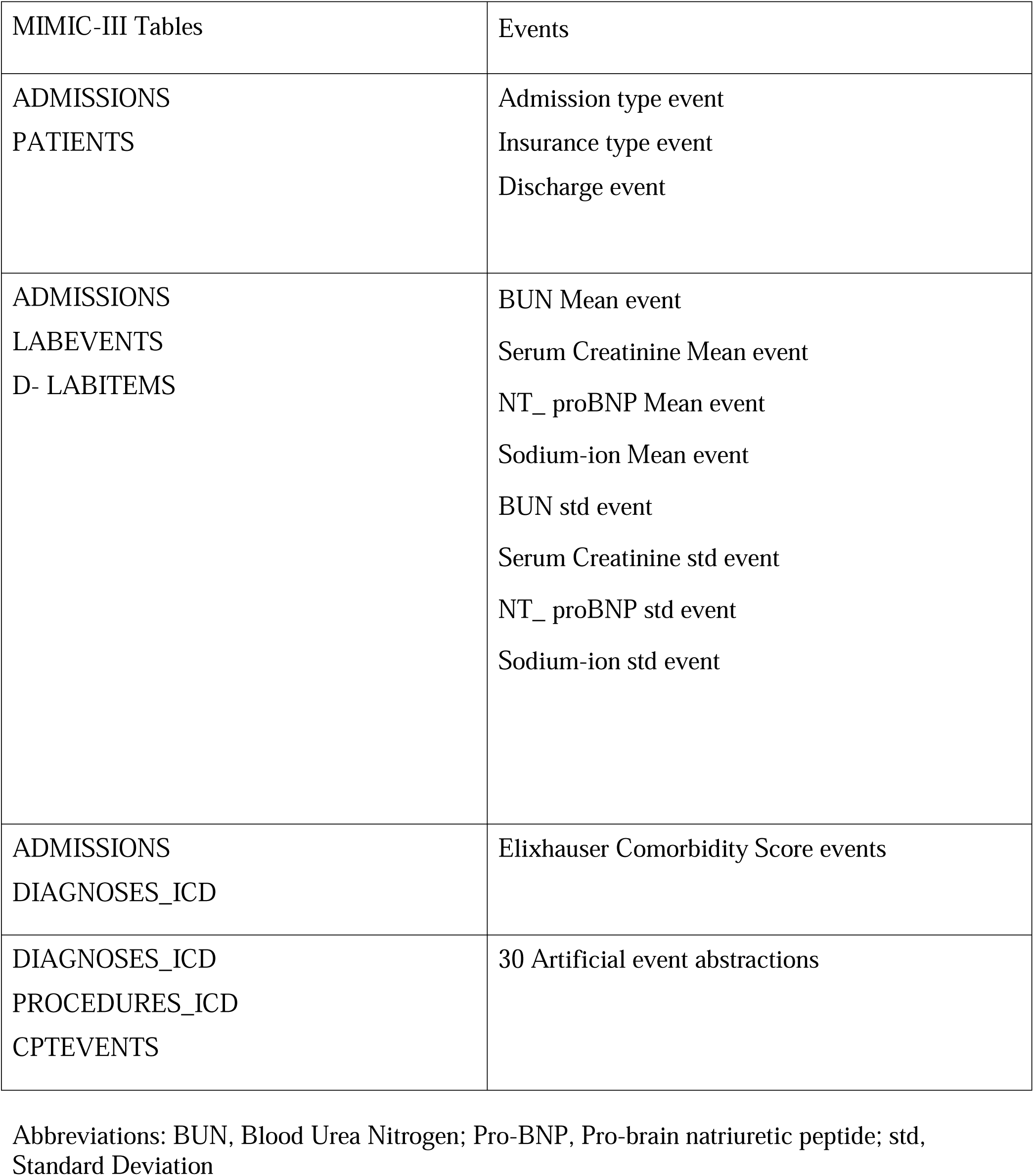
Mapping of the medical health records of the patients from the MIMIC-III database to the event logs

### Event Types and the Associated Timestamps

For each patient, we converted the EHR to events with the following sequence: First, we considered the admission type event with the admission time as its timestamp. Admission type event was important since it distinguished whether the admission was planned or unplanned. The second event was the insurance type event with a timestamp of 1ms after the timestamp of the admission type to maintain the order of the events. Insurance type was chosen since it could possibly affect the discharge/transfer rate.

The next set of events were the lab measurements. Specific HF-related lab measurements were chosen based on the literature [11,12] and experts’ opinions. The lab measurements might be measured once or several times for each patient. Two types of events were created for each lab item, out of which one was the Mean of the specific lab item and the other was the Standard Deviation (std) of each lab item.

In the cases in which the lab item was measured once, the timestamp of the Mean event was set to the timestamp at which the lab item was measured initially. The timestamp for the std event was set to 1ms after the timestamp of the Mean event. For the lab measurements, which were measured several times, we performed similarly for the Mean event. However, for the std event of such cases, the timestamp was set to the last time at which the lab item was measured. We considered a separate set of events representing the comorbidities. Elixhauser comorbidity score was calculated by using the ICD-9 diagnosis codes for each hospital visit [19]. A specific comorbidity group can be determined by assigning points through the Elixhauser comorbidity score if particular ICD-9 codes are present. These comorbidity events were created because they represent what diseases have been diagnosed, whether the diseases are chronic, and the criticality of the patients. Additionally, based on the literature, these events were strongly associated with ICU readmission risk [20]. In cases where a point was assigned to a specific group, an event was created with the same name as in the comorbidity group. Since the focus of this paper was on readmission prediction, we needed specific event logs that separated all the timestamps of the events from the discharge timestamp (which was the final event). Therefore, the timestamps for these events were set to be very close to the discharge time of the relevant hospital visit. In cases where multiple comorbidity events were created, the timestamp for the second comorbidity event was set 1ms after the timestamp of the first event. The same logic was applied for the next comorbidity events as well.

The artificial events were created from the sequence of CPT, and ICD-9-CM observations codes as by Theis et al. [16]. We considered these events since they represented the diagnoses and procedures of a patient which were likely to be important factors to predict readmission. The artificial events’ timestamps were set to the timestamps of the sequence of the observations plus 1ms. These timestamps accordingly were compared with timestamps of the discharge event, and they were set to a time before the discharge timestamp to ensure the orders of the events were maintained.

In the end, the discharge event was created for each hospital admission and the timestamp of this event was set to the discharge time of the patient for the corresponding admission. This event was created since this was a point at which the next event (admission type event) would be predicted by using Decay Replay Mining (DREAM) algorithm [21].

Note that the addition of 1ms to the timestamps in our conversion did not alter any information since the time dimension in MIMIC-III was days and subsequently negligible in our analysis.

### Unplanned 30-day Readmission Prediction

We proposed a process mining approach for unplanned 30-day readmission prediction. The resultant event logs were fed to the DREAM algorithm to generate the time information (TSS). The severity scores on admission day including the Charlson [22] and Elixhauser scores were used as independent variables. Charlson score method assigns higher weights to more severe and critical conditions as compared to Elixhauser that assigns the same weight to all conditions. The generated TSS, together with the demographic information and the severity scores were then fed to a Neural Network (NN) model to predict unplanned 30-day readmission of the ICU patients with HF. Figure 1 illustrates the overview of the proposed model. The corresponding source code is publicly available on our Github repository.

**Figure 1.**
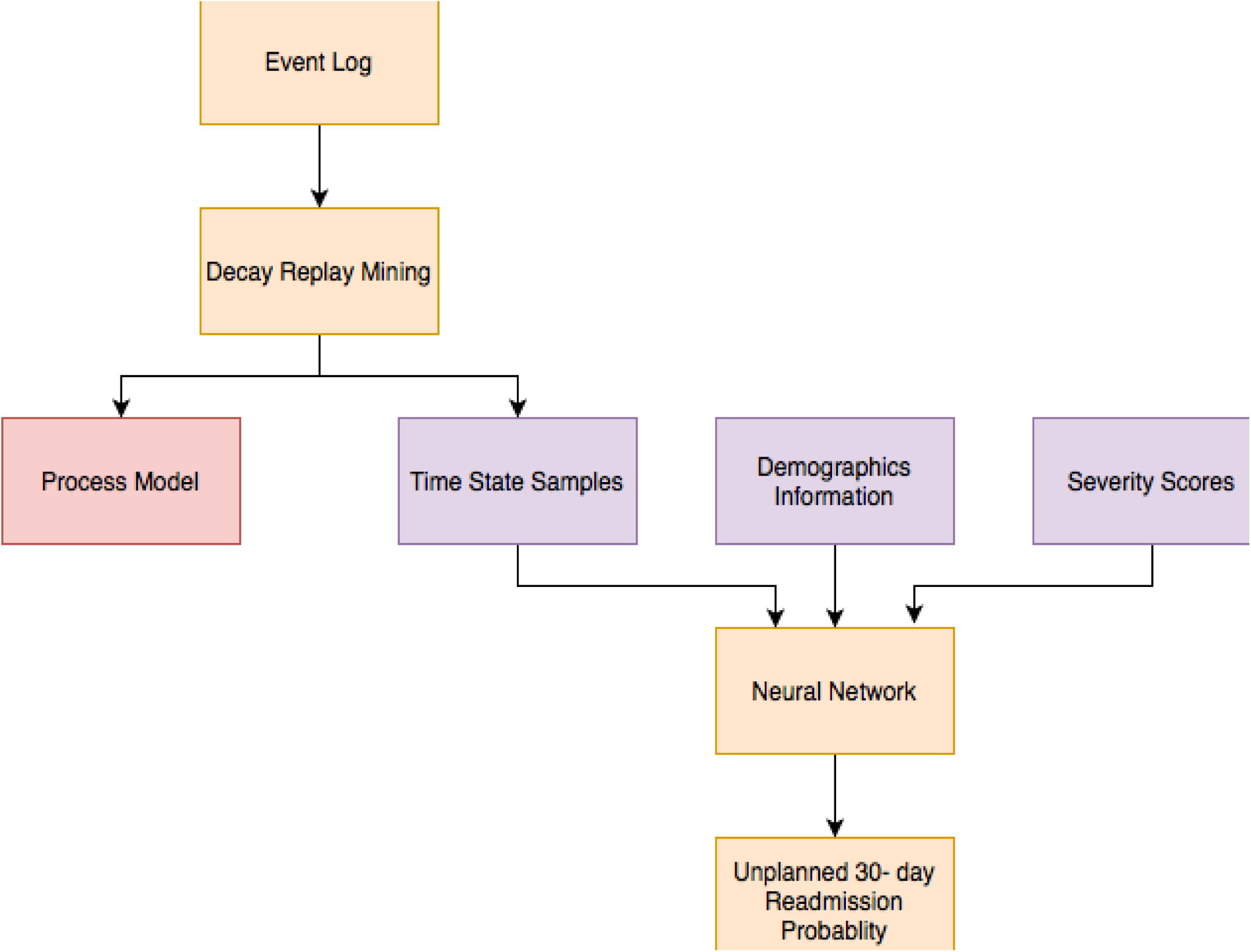
Overview of the methodology

The proposed model was evaluated by calculating Area Under the Receiver Operating Characteristic curve (AUROC), precision, sensitivity, and F-score on the test set. To obtain 95% Confidence Intervals (CIs) of the AUROC value, DeLong’s method was used [23].

### Statistical Analysis between Cohorts

The derivation and validation cohorts were compared using Chi-Square and two-sided t-tests. For the comparison of the categorical variables, Chi-Square tests were performed, and for continuous variables, t-tests were implied. The significant level was determined based on P < 0.05. Descriptive statistics, model development, and statistical analysis were conducted using Python, version 3.6.

### Variables Impacts

Shapley value analysis [24] was conducted on the test set to find out the impact of each variable in our model prediction and to figure out which variable was particularly associated with readmissions. The Shapley values described the Mean contribution of each variable to the outcome across different coalitions [16].

## RESULTS

### Cohort Characteristics Model Completion

Following the approach for selection of HF patients discussed before in this paper, a subset of 3411 patients was selected from the MIMIC-III database. The selected cohort was then split into derivation and test sets randomly with a ratio of 84/16, which yielded a result of 2856 patients for derivation and 555 patients for the test cohorts. Moreover, the derivation cohort was further split into derivation and validation with the ratio of 85/15 resulting in 2422 patients for derivation and the remaining 434 patients for validation cohorts, to be used in the process discovery step and the NN derivation. Lastly, the best model was chosen for further evaluation on the test cohort. The description of the derivation and validation cohorts is presented in Table 2. In terms of age, the validation cohort (70.4 years) was slightly older than the derivation cohort (69.9 years) with a P of 0.228 which showed there were no significant differences between cohorts. In terms of gender, the derivation cohort (47.6 %) contained slightly more females compared to the validation cohort (46.3 %). The whole distribution of the race was not significantly different between the cohorts [P = 0.270], of which the details are shown in Table 2. The proportions of white patients in the derivation and validation cohorts were 75.8 %, and 74.9 % respectively. The lab measurements were not significantly different between cohorts except for Urea Nitrogen which was 0.017.

**Table 2:** Comparison of the variables including outcome, demographics, and laboratory findings between derivation and validation cohorts

| Characteristics | Derivation Cohort (N = 2422) | Validation Cohort (N= 434) | P |
| --- | --- | --- | --- |
| Outcome Variable N, (%) |  |  |  |
| Readmission | 581 (23.9) | 102 (23.5) | 0.270 |
| Demographics |  |  |  |
| Age Mean (std) | 69.9 (14.3) | 70.4 (13.9) | 0.228 |
| Female (%) | 47.6 | 46.3 | 1.00 |
| Race N (%) |  |  | 0.270 |
| African American | 291 (12.0) | 51 (11.8) |  |
| Hispanic | 76 (3.10) | 18 (4.15) |  |
| Others, non- Hispanic | 170 (7.00) | 37 (8.53) |  |
| White | 1835 (75.8) | 325 (74.9) |  |
| Asian | 50 (2.10) | 3 (0.691) |  |
| Laboratory Findings Mean (std) |  |  |  |
| Sodium | 138.6 (4.58) | 138.5 (4.80) | 0.835 |
| Urea Nitrogen | 34.4 (24.0) | 32.6 (22.3) | 0.017 |
| NT proBNP | 0.187 (0.380) | 0.181 (0.385) | 0.613 |
| Serum Creatinine | 0.002 (0.04955) | 0.004 (0.062) | 0.083 |

Abbreviations: Pro-BNP, Pro-brain natriuretic peptide

### Shapley Value Analysis

Figure 2 illustrates the results of the Shapley value analysis. Based on this figure, severity scores had the most significant impact on the prediction of unplanned 30-day readmission of the HF ICU patients, followed by demographic information and admission events that seemed to have a similar impact on prediction. Whereas artificial events, comorbidity events, and lab measurement events were the least important variables for the prediction of the outcome in order. Among the severity scores, Charlson had a higher impact on prediction compared to that of Elixhauser which showed that the severity of the conditions played an important role in the prediction of unplanned 30-day readmission of ICU HF patients since Charlson score assigns higher weights to the severity level of the conditions than Elixhauser.

**Figure 2.**
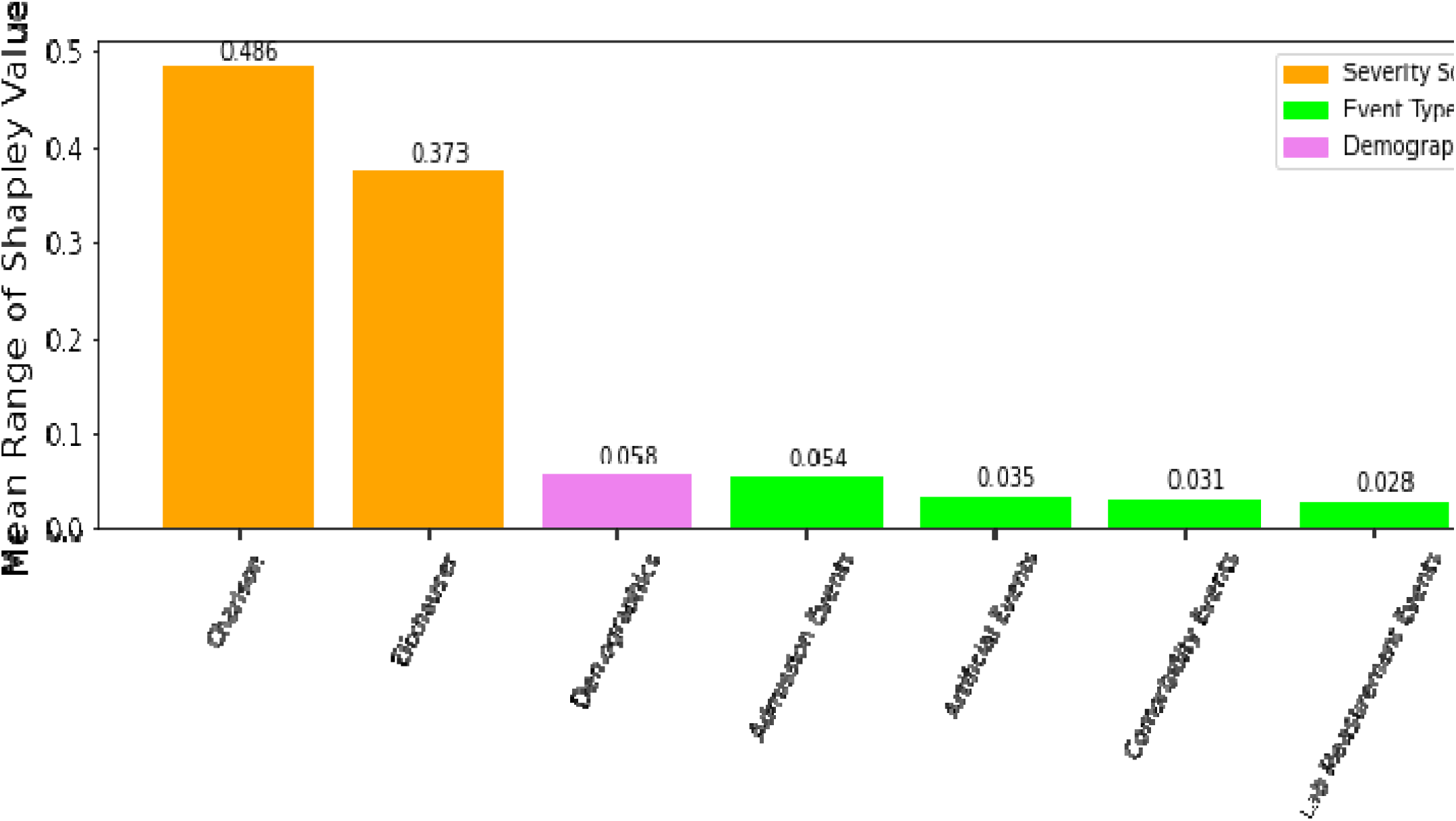
The Mean range of Shapley Values for each variable type

The Shapley value analysis confirmed that the severity scores had the highest impact on prediction in our model. However, other contributing factors impacted the prediction of the outcome including demographics, admission events, artificial events, comorbidity events, and lab measurement events which were all ignored by health calculators as the inputs for prediction.

### Evaluation Metrics and Proposed Model Performance

The proposed approach resulted in the following metrics, AUROC, 95% CI, precision, sensitivity, and F-score are as follows, respectively: 0.920, CI of [0.891-0.962], 0.880, 0.781, 0.800.

## DISCUSSION

### Existing Model Compilation Summary

Several methods have been concurrently developed to predict unplanned 30-day readmission of the ICU patients with HF aiming to benefit both health care providers and the patients. Table 3 shows the existing models which have been developed to predict unplanned 30-day readmission of ICU patients with HF by using the MIMIC-III dataset.

**Table 3:**
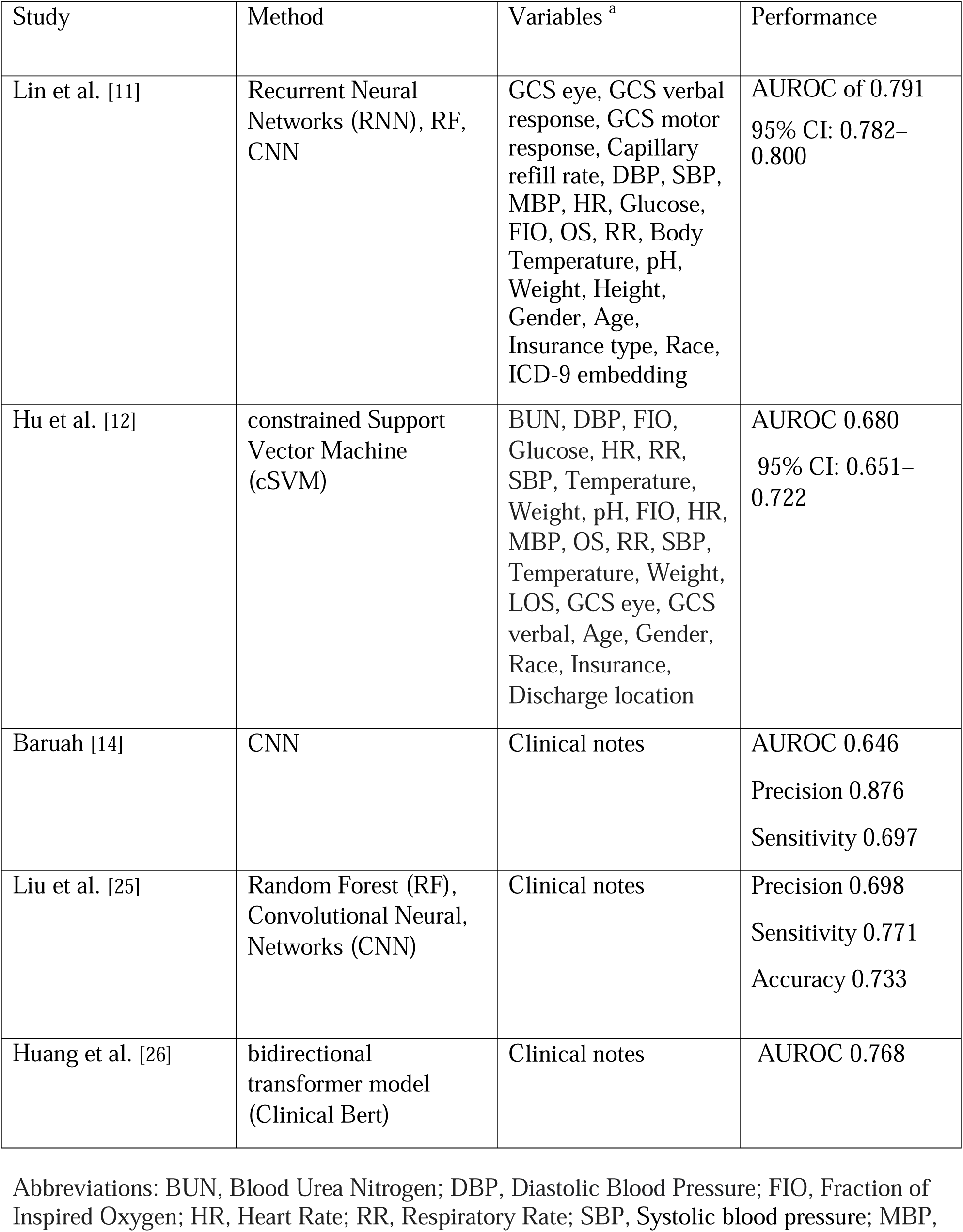
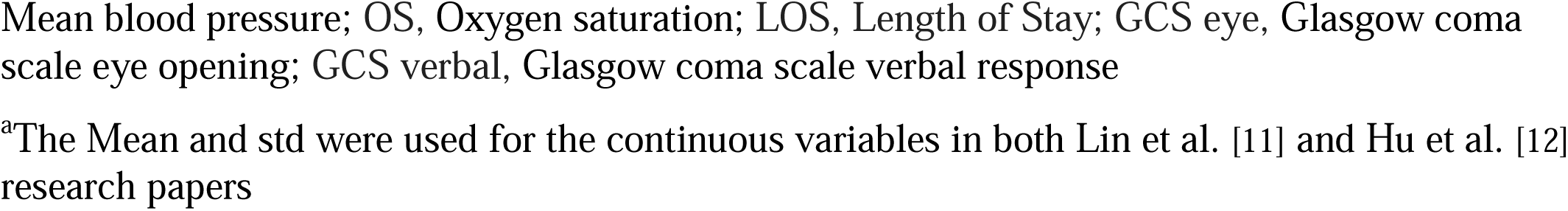
Summary of the existing models and their performance on the MIMIC-III dataset

| Study | Method | Variables <sup>a</sup> | Performance |
| --- | --- | --- | --- |
| Lin et al. [11] | Recurrent Neural Networks (RNN), RF, CNN | GCS eye, GCS verbal response, GCS motor response, Capillary refill rate, DBP, SBP, MBP, HR, Glucose, FIO, OS, RR, Body Temperature, pH, Weight, Height, Gender, Age, Insurance type, Race, ICD-9 embedding | AUROC of 0.791<br>95% CI: 0.782–0.800 |
| Hu et al. [12] | constrained Support Vector Machine (cSVM) | BUN, DBP, FIO, Glucose, HR, RR, SBP, Temperature, Weight, pH, FIO, HR, MBP, OS, RR, SBP, Temperature, Weight, LOS, GCS eye, GCS verbal, Age, Gender, Race, Insurance, Discharge location | AUROC 0.680<br>95% CI: 0.651–0.722 |
| Baruah [14] | CNN | Clinical notes | AUROC 0.646<br>Precision 0.876<br>Sensitivity 0.697 |
| Liu et al. [25] | Random Forest (RF), Convolutional Neural Networks (CNN) | Clinical notes | Precision 0.698<br>Sensitivity 0.771<br>Accuracy 0.733 |
| Huang et al. [26] | bidirectional transformer model (Clinical Bert) | Clinical notes | AUROC 0.768 |

Abbreviations: BUN, Blood Urea Nitrogen; DBP, Diastolic Blood Pressure; FIO, Fraction of Inspired Oxygen; HR, Heart Rate; RR, Respiratory Rate; SBP, Systolic blood pressure; MBP,
Mean blood pressure; OS, Oxygen saturation; LOS, Length of Stay; GCS eye, Glasgow coma scale eye opening; GCS verbal, Glasgow coma scale verbal response
<sup>a</sup>The Mean and std were used for the continuous variables in both Lin et al. [11] and Hu et al. [12] research papers

In this study, a process mining technique was investigated for predicting unplanned 30-day readmission of ICU patients with HF, in which time information associated with the events, severity scores, and demographics were fed into a NN model.

The effectiveness of our developed approach outperformed the best results of the existing literature in terms of the AUROC value proposed by Lin et al [11]. The efficacy of our approach was demonstrated by a substantial improvement of +10% on AUROC.

In addition, the presented results indicated +6% and +7% improvements in sensitivity and F-score metrics, respectively, compared to the best sensitivity and F-score values reported in the literature by Liu et al [25].

Although the existing proposed methodologies in the literature were successful in predicting unplanned 30-day readmission of ICU patients with HF, they possessed several drawbacks. First, most of the existing models did not use the time-series features, and to the best of our knowledge, none of them incorporated time information associated with the variables in their predictive modeling that could lead to significant information loss and poor performance accordingly [26,27].

Furthermore, the proposed approach was a process mining approach that illustrated the *careflows* of patients through a process model. As a result, our framework was more interpretable compared to the existing methods, which is significant for clinical applications [28].

Moreover, the health calculators that computed outcomes based on the severity scores ignored the past medical history of the patients which could have a significant impact on the likelihood of unplanned readmission.

Our proposed approach had several advantages over prior research papers which are as follows: a) Process mining approach yielded a comprehensive analysis of *careflows* of patients through a process model which is understandable and can easily be interpreted compared to ML techniques. The process model provided a map that represented the possible diagnosis, procedures, performed services, laboratory measurements, and more, that happened to a patient.

Additionally, it eased the interpretability of a model prediction. An example of a process model can be found in the existing research paper [15,29] b) The EHR can be directly used as inputs to our proposed approach without any computationally expensive preprocessing steps. c) The process mining framework was capable of modeling the time-related variables and incorporating the medical history of the patients from the previous hospital visits in the prediction algorithm unlike ML-based models and health calculators.

### Study Limitations

The proposed approach had some limitations. Since this approach was a process mining approach, the availability of the past hospital visits of the patients was essential. This approach was not useful for patients whose admission histories were not available. However, this limitation can be overcome if the history of patients could be exchanged through a network system between health care providers. Application Program Interfaces (APIs) and similar innovations hold promise that soon these drawbacks can seemingly be curtailed.

Moreover, in our model development, the derivation and validation datasets were used to build the model. The test dataset was set aside from the beginning and only used to evaluate the performance of the model. The derivation, validation, and test sets were coming from the MIMIC-III dataset. However, using an independent dataset from a different system would be beneficial to test the performance of the model [30], which provides room for future work.

## CONCLUSIONS

A process mining approach to model EHR data of ICU patients with HF to predict unplanned 30-day readmission provided significant improvement in outcome prediction observed and compared to the results of the existing literature. This improvement could be due to the capability of the process mining approach of modeling time information related to the variables and incorporating past hospital visits of the patients for prediction. Our framework can assist clinicians in identifying patients with a higher risk of unplanned 30-day readmission. Discharge planners may find this prediction tool useful in determining when a patient is safe to be discharged from the hospital and to guide post-discharge outpatient management. Future studies may validate the proposed approach using datasets from other healthcare systems or investigate its use for different diseases and outcomes. Moreover, the MIMIC-III dataset contains useful information such as clinical notes, and images, which can be fed to the models as inputs. Therefore, it potentially makes room for further research.

## Data Availability

Data is publically available

https://physionet.org/content/mimiciii-demo/1.4/

## ABBREVIATIONS

DREAM: Decay Replay Mining;
NN: Neural Network;
TSS: Time State Sample;
BUN: Blood Urea Nitrogen;
MIMIC-III: Medical Information Mart for Intensive Care III;
std: Standard Deviation;
Pro-BNP: Pro-brain natriuretic peptide.

## DECLARATIONS

### Ethics approval and consent to participate

Not applicable.

### Consent for publication

Not applicable.

### Availability of data and materials

The MIMIC-III database which were used during the current study are publicly available.

### Authors’ contributions

M.P. and H.D.: Involved in all aspects of this study. J.T.: Data acquisition, data interpretation, drafting of the manuscript. M.R. and A.A.: Data interpretation and drafting of the initial manuscript. H.A.: drafting of the initial manuscript.

## Acknowledgements

The authors would also like to thank the developers of MIMIC III for providing a detailed and comprehensive public EHR dataset.

## REFERENCES

1. Mozaffarian D BE, Go A, Arnett D, Blaha M, Cushman M, et al. Heart disease and stroke statistics-2016 update: a report from the American Heart Association. Circulation.; 2016.

2. Golas SB, Shibahara T, Agboola S, Otaki H, Sato J, Nakae T, et al. A machine learning model to predict the risk of 30-day readmissions in patients with heart failure: a retrospective analysis of electronic medical records data. BMC Medical Informatics and Decision Making. 2018;18(1):44

3. Tan B-y, Gu J-y, Wei H-y, Chen L, Yan S-l, Deng N. Electronic medical record-based model to predict the risk of 90-day readmission for patients with heart failure. BMC Medical Informatics and Decision Making. 2019;19(1):193.

4. Ponzoni CR, Corrêa TA-O, Filho RR, Serpa Neto A, Assunção MSC, Pardini A, et al. Readmission to the Intensive Care Unit: Incidence, Risk Factors, Resource Use, and Outcomes. A Retrospective Cohort Study. (2325-6621 (Electronic)).

5. Desautels T, Das R, Calvert J, Trivedi M, Summers C, Wales DJ, et al. Prediction of early unplanned intensive care unit readmission in a UK tertiary care hospital: a cross-sectional machine learning approach. BMJ Open. 2017;7(9):e017199.

6. Singer De Fau - Mulley AG, Mulley Ag Fau - Thibault GE, Thibault Ge Fau - Barnett GO, Barnett GO. Unexpected readmissions to the coronary-care unit during recovery from acute myocardial infarction. (0028-4793 (Print)).

7. Ashfaq A, Sant’Anna A, Lingman M, Nowaczyk S. Readmission prediction using deep learning on electronic health records. Journal of Biomedical Informatics. 2019;97:103256.

8. Hua M, Gong Mn Fau - Brady J, Brady J Fau - Wunsch H, Wunsch H. Early and late unplanned rehospitalizations for survivors of critical illness*. (1530-0293 (Electronic)).

9. Vader JM, LaRue SJ, Stevens SR, Mentz RJ, DeVore AD, Lala A, et al. Timing and Causes of Readmission After Acute Heart Failure Hospitalization-Insights From the Heart Failure Network Trials. (1532-8414 (Electronic)).

10. Miotto R, Li L, Kidd BA, Dudley JT. Deep Patient: An Unsupervised Representation to Predict the Future of Patients from the Electronic Health Records.Scientific Reports. 2016;6(1):26094.

11. Lin YW, Zhou Y, Faghri FA-O, Shaw MJ, Campbell RH. Analysis and prediction of unplanned intensive care unit readmission using recurrent neural networks with long short-term memory. (1932-6203 (Electronic)).

12. Zhiyong Hu DD. A new analytical framework for missing data imputation and classification with uncertainty: Missing data imputation and heart failure readmission prediction. PLoS One. 2020.

13. Ghasemi M, Amyot D. Process mining in healthcare: A systematised literature review. International Journal of Electronic Healthcare. 2016;9:60.

14. Baruah P. Predicting Hospital Readmission using Unstructured Clinical Note 2020.

15. Pishgar M RM, Theis J, Darabi H. Process Mining Model to Predict Mortality in Paralytic Ileus Patients. International Conference on Cyber-physical Social Intelligence 2021.

16. Theis J Fau - Galanter W, Galanter W Fau - Boyd A, Boyd A Fau - Darabi H, Darabi H. Improving the In-Hospital Mortality Prediction of Diabetes ICU Patients Using a Process Mining/Deep Learning Architecture. LID - 10.1109/JBHI.2021.3092969 [doi]. (2168-2208 (Electronic)).

17. Johnson AEaP, Tom J and Shen, Lu and Lehman Li{-}wei H and Feng, Mengling and Ghassemi, Mohammad and Moody, Benjamin and Szolovits, Peter and Celi Leo Anthony and Mark Roger G. MIMIC-III, a freely accessible critical care database. Scientific data. 2016;3:160035.

18. Rojas E, Munoz-Gama J, Sepúlveda M, Capurro D. Process mining in healthcare: A literature review. Journal of Biomedical Informatics. 2016;61:224–36.

19. Elixhauser A, Steiner C Fau - Harris DR, Harris Dr Fau - Coffey RM, Coffey RM. Comorbidity measures for use with administrative data. (0025-7079 (Print)).

20. Brown SE, Ratcliffe Sj Fau - Halpern SD, Halpern SD. An empirical derivation of the optimal time interval for defining ICU readmissions. (1537-1948 (Electronic)).

21. Theis J, Darabi H. Decay Replay Mining to Predict Next Process Events. IEEE access : practical innovations, open solutions. 2019;7:119787–803.

22. Charlson Me Fau - Pompei P, Pompei P Fau - Ales KL, Ales Kl Fau - MacKenzie CR, MacKenzie CR. A new method of classifying prognostic comorbidity in longitudinal studies: development and validation. (0021-9681 (Print)).

23. DeLong ER, DeLong Dm Fau - Clarke-Pearson DL, Clarke-Pearson DL. Comparing the areas under two or more correlated receiver operating characteristic curves: a nonparametric approach. (0006-341X (Print)).

24. S. M. Lundberg and S.-I. Lee. A unified approach to interpreting model predictions. 2017.

25. Liu X, Chen, Y., Bae, J., Li, H., Johnston, J., & Sanger, T. Predicting Heart Failure Readmission from Clinical Notes Using Deep Learning. IEEE International Conference on Bioinformatics and Biomedicine (BIBM).

26. Huang K AJ, Ranganath R. ClinicalBERT: Modeling Clinical Notes and Predicting Hospital Readmission. 2020.

27. Feng M, McSparron JI, Kien DT, Stone DJ, Roberts DH, Schwartzstein RM, et al. Transthoracic echocardiography and mortality in sepsis: analysis of the MIMIC-III database. (1432-1238 (Electronic)).

28. Kurniati AA-O, Rojas E, Hogg D, Hall G, Johnson OA. The assessment of data quality issues for process mining in healthcare using Medical Information Mart for Intensive Care III, a freely available e-health record database. (1741-2811 (Electronic)).

29. Darabi H, Galanter WL, Lin JY, Buy U, Sampath R, editors. Modeling and integration of hospital information systems with Petri nets. 2009 IEEE/INFORMS International Conference on Service Operations, Logistics and Informatics; 2009 22–24 July 2009.

30. Wessler BS, Nelson J, Park JG, McGinnes H, Gulati G, Brazil R, et al. External Validations of Cardiovascular Clinical Prediction Models: A Large-Scale Review of the Literature. Circulation: Cardiovascular Quality and Outcomes.0(0):CIRCOUTCOMES.121.007858.

